# A Modified Thromboelastometry Assay Enables Rapid, Real-Time Evaluation of Complement-Driven Immunothrombosis

**DOI:** 10.64898/2026.06.30.26356356

**Authors:** Evangelos Papadimitriou, Anastasia-Maria Natsi, Charalampos Papagoras, Dimitrios Mastellos, Victoria Tsironidou, Ioannis Mitroulis, John D. Lambris, Konstantinos Ritis

**Affiliations:** First Department of Internal Medicine and Laboratory of Molecular Hematology, University Hospital of Alexandroupolis, Democritus University of Thrace, Alexandroupolis, Greece; National Center for Scientific Research ‘Demokritos’, Athens, Greece; Department of Pathology and Laboratory Medicine, Perelman School of Medicine, University of Pennsylvania, Philadelphia, USA

**Keywords:** Immunothrombosis, complement system, C3, Cp40, thromboelastometry, COVID-19, Antiphospholipid syndrome, Rheumatoid arthritis

## Abstract

**Introduction:** Complement and coagulation are tightly interconnected systems that contribute to immunothrombosis and can drive inflammatory or thrombotic diseases. Leveraging this relationship and crosstalk we developed a method to functionally evaluate complement-induced coagulation activity using thromboelastometry (thermoelastometry of complement-driven immunothrombosis; TCDI).

**Methods:** To study the complement-dependent activation of coagulation, platelet-poor plasma (PPP) from patients was mixed with healthy blood in the presence or absence of the compstatin-based C3 inhibitor Cp40. PPP from healthy controls (n=10), or from patients with antiphospholipid syndrome (APS; n=6), severe COVID-19 (n=13), rheumatoid arthritis (RA; n=7), or synovial fluid (SF) from RA patients, were analyzed for their capacity to induce complement activation in healthy blood. Whole blood coagulation was analyzed by thromboelastometry and complement-driven immunothrombosis was quantified as clotting time (CT) prolongation following Cp40 treatment, expressed as fractional difference percentage (FD%). In parallel, C3a generation was measured by ELISA to monitor the C3 inhibitory activity of Cp40.

**Results:** Plasma from patients with APS and COVID-19 induced significant CT prolongation following C3 inhibition by Cp40 and increased FD% values compared with controls, indicating active complement-driven immunothrombosis. Higher TCDI levels were associated with mortality in severe COVID-19. In RA, TCDI positivity was detected in synovial fluid (SF) rather than peripheral plasma. Moreover, TCDI-positive samples treated with Cp40 exhibited significant inhibition of C3a generation, which strongly correlated with FD% values (r=0.67, p=0.0005).

**Conclusion:** The TCDI assay may provide a rapid, real-time evaluation of immunothrombotic activity in inflammatory and thrombotic disorders, which could inform timely medical prevention.

## Introduction

The proteolytic pathways of the complement and coagulation systems both contribute to protecting the host from injury and infection through tightly coordinated inflammatory and hemostatic responses^1^. This is accomplished by the functional bridging of innate immunity and coagulation in a process recently defined as immunothrombosis^2^. Coagulation and complement are evolutionarily conserved systems that serve essential roles in hemostasis and host defense, respectively. However, their dysregulated activation and crosstalk can promote immunothrombosis, triggering excessive inflammatory and prothrombotic responses through multiple humoral and cellular signaling cascades, including those driven by coagulation proteases^3,4^.

Among the key mediators of innate immunity, complement and neutrophils are central to the pathophysiology and clinical phenotype of diseases such as COVID-19 and antiphospholipid syndrome (APS)^5–7^. Neutrophil extracellular traps (NETs) constitute an established pathophysiological link between complement activation and coagulation, serving as a source of active tissue factor (TF) either through direct surface expression or via TF-positive extracellular vesicles. Moreover, their negatively charged components activate the contact pathway of coagulation, promoting the generation of plasma kallikrein, FXIIa, and FXIa^8–12^.

Despite extensive mechanistic insights into immunothrombosis, there is currently no real-time functional assay capable of identifying its upstream triggers or distinguishing between systemic and local activation. Thromboelastometry (TEM) provides clinically relevant coagulation parameters that reflect thrombotic phenotypes. Thus, a modified TEM-based approach detecting complement-driven immunothrombosis would enable a functional and clinically relevant assessment across diverse diseases.

We therefore optimized a TEM-based assay to link complement activation with thrombotic and inflammatory manifestations. This assay, termed thromboelastometry of complement-driven immunothrombosis (TCDI), was developed based on our previous experimental and clinical observations in COVID-19 and APS patients, showing that systemic complement activation drives immunothrombosis in these disorders. Specifically, plasma from affected patients activates complement in healthy donor blood, thereby triggering inflammatory and thrombotic responses, including activation of monocytes, platelets, and neutrophils through NET formation^3,5,7, 10, 13, 14^.

## Materials and Methods

### 1. Patients and sampling

Platelet-poor plasma (PPP) from 13 patients with severe COVID-19 (PaO_2_/FiO_2_ <180) and 6 patients with APS (**Tables 1 and 2**) were analyzed using the TCDI assay, alongside plasma from 10 healthy donors as controls (**Supplementary Table 1**). Rheumatoid arthritis (RA) was included as a control disease model. To investigate whether the TCDI observations apply to the systemic or local/intra-articular level of inflammation, we recruited patients with RA exacerbation and synovial effusion in at least one knee joint. Paired samples of peripheral blood and knee synovial fluid (SF) aspirates were obtained from all RA patients (n=7, **Table 3**).

**Table 1.** Characteristics of Covid-19 patients. FD, fractional difference; M, Male; F, Female; P/F ratio, the ratio of the arterial partial pressure of oxygen (PaO□) to the fraction of inhaled oxygen (FiO□), used to assess the efficiency of oxygen transfer from the lungs to the blood; N/A, not applicable; Marked as N/A because the data point was unavailable. **†** These samples were available for C3a ELISA.

| Patient ID | Age | Gender | FD | P/F ratio | Survived |
| --- | --- | --- | --- | --- | --- |
| **Covid 1** | 50-59 | M | 14.1 | N/A | No |
| †Covid 2 | 50-59 | M | 4.5 | 77 | Yes |
| **Covid 3** | 50-59 | F | 3.6 | 179.24 | Yes |
| **Covid 4** | 50-59 | M | 2.9 | 173.02 | Yes |
| †Covid 5 | 40-49 | M | 9.9 | 105 | Yes |
| †Covid 6 | 40-49 | M | 7.1 | 73.3 | No |
| †Covid 7 | 30-39 | M | 1.6 | 123.64 | Yes |
| **Covid 8** | 40-49 | M | 6.8 | 172.3 | Yes |
| †Covid 9 | 70-79 | M | 8 | N/A | No |
| **Covid 10** | 60-69 | M | 3.1 | 103 | Yes |
| †Covid 11 | 60-69 | M | 2.2 | 178.33 | No |
| †Covid 12 | 60-69 | M | 6.4 | 66.67 | No |
| †Covid 13 | 70-79 | M | 13.8 | 101.11 | No |

**Table 2.** Characteristics of antiphospholipid syndrome (APS) patients. FD, Fractional Difference; M, male; F, female. Patients with APS had confirmed antiphospholipid antibody (aPL) positivity on at least two occasions 12 weeks apart, according to the APS classification criteria ^6^. * For these patients, additional to an early aPL-positive plasma sample, another aPL-negative sample obtained at the 3-year follow-up (T2) was available. The aPL-positive sample rendered a positive TCDI assay, while the aPL-negative did not. **†** These samples were available for C3a ELISA.

| **Patient ID** | **Age** | **Gender** | **FD** | **Antibody positive\*** |
| --- | --- | --- | --- | --- |
| † APS 1 | 40-49 | F | 6.12 | + |
| † APS 1 T2 | 40-49 | F | 0 | - |
| APS 2 | 50-59 | F | 6.61 | + |
| † APS 3 | 40-49 | M | 2.8 | + |
| † APS 4 | 50-59 | F | 7.01 | + |
| † APS 4 T2 | 50-59 | F | 0 | - |
| APS 5 | 50-59 | F | 4.53 | + |
| † APS 6 | 40-49 | M | 4.35 | + |

**Table 3.** Characteristics of Rheumatoid arthritis patients. RA, rheumatoid arthritis; DAS28, Disease Activity Score based on 28 joints; higher scores indicate greater rheumatoid arthritis activity; RF, Rheumatoid Factor; CCP, cyclic citrullinated peptide antibodies; SF, synovial fluid; PPP, platelet-poor plasma; FD, fractional difference. * TCDI analysis of citrated plasma yielded a positive result, consistent with systemic involvement, as evidenced by the presence of fever, polyarthritis, elevated inflammatory markers, and high disease activity according to the DAS28 score. **†** These samples were available for C3a ELISA.

| **Patient ID** | **Age** | **Gender** | **DAS28** | **Antibody Positivity** | **SF FD** | **PPP FD** |
| --- | --- | --- | --- | --- | --- | --- |
| †RA 1 | 60-69 | F | 3.95 | RF(-), CCP(+) | 10.08 | 0 |
| RA 2 | 60-69 | F | 4.12 | RF(-), CCP(+) | 3.26 | 0 |
| †RA 3 | 80-89 | F | 5.19 | RF(+), CCP(+) | 3.21 | 0 |
| RA 4 | 60-69 | M | 3.95 | RF(-), CCP(+) | 1.58 | 0 |
| RA 5* | 40-49 | F | 5.66 | RF(-), CCP(+) | 8.26 | 9.2 |
| †RA 6 | 40-49 | F | 5.49 | RF(-), CCP(+) | 4.3 | 0 |
| †RA 7 | 60-69 | F | 4.8 | RF(+), CCP(+) | 2.5 | 0 |

All patients provided written informed consent prior to sample collection. Patients with APS and RA were recruited from the First Department of Internal Medicine, University Hospital of Alexandroupolis, and from the Rheumatology Outpatient Clinic. Patients with COVID-19 were participants in the ITHAKA study, as previously described^13^. Healthy donors were recruited as voluntary participants. The protocol for the present study was approved by the Ethics Committee of the University Hospital of Alexandroupolis.

### 2. Thromboelastometry of complement-driven immunothrombosis (TCDI)

Peripheral venous blood was collected from healthy donors into sodium citrate tubes (PC Vacutainer®, BD Biosciences) and processed within 1 hour of collection. Only samples with platelet counts within the normal range (150–350 ×10^3^/μL) were used.

For each experiment, 300 μL of citrated whole blood from healthy donors was incubated with 100 μL of PPP collected from patients with COVID-19, APS, RA, or from healthy donors. For experiments involving SF from RA patients, SF was added to healthy donor whole blood at a final concentration of 2.5% (v/v; prepared as a 1:10 dilution). Samples were prewarmed for 10 minutes at 37°C prior to their mixing with donor blood.

In parallel, to evaluate the contribution of complement activation to coagulation, 100 μL of prewarmed PPP from patients or controls or diluted SF samples (diluted 1:10 in PBS) were pre-incubated (30 minutes at 37°C) with the compstatin-based C3 inhibitor Cp40 (clinically developed as AMY-101; Amyndas Pharmaceuticals) at a concentration of 20 μM. The pretreated samples were then mixed with 300 μL of citrated whole blood and incubated for an additional 30 minutes at 37°C. Cp40 was added to maintain a final concentration of 20 μM in the complete reaction mixture. In selected experiments, instead of Cp40, DNase I (1 U/mL; Thermo Fisher Scientific) was used as an alternative intervention to dismantle NETs under similar experimental conditions.

Following incubation, 350 μL out of 400 μL of the final mixture were transferred to pre-warmed ClotPro cups. Whole blood coagulation was assessed using the ClotPro® viscoelastic coagulation analyzer (enicor GmbH, Germany). Coagulation was initiated through the addition of 20 μL of freshly prepared 0.2M anhydrous CaCl□, (Merck Millipore; 0.111 g in 5 mL sterile distilled water), pre-warmed to 37°C. Strict temperature control (36.5–37°C) and freshly prepared CaCl□ were maintained throughout to ensure reproducibility.

Samples were analyzed according to the manufacturer’s instructions. Coagulation parameters, including clotting time (CT), clot formation time (CFT), and maximum clot firmness (MCF) were recorded. CT values were recorded in seconds. For comparative analysis, samples that yielded a zero or negative fractional difference, indicating an absence of CT prolongation relative to the control, were classified as negative and assigned a value of 0.

Although clot formation time (CFT) and maximum clot firmness (MCF) were also assessed, no significant differences were observed between experimental conditions. Therefore, only CT values were included in the final analysis.

All experiments were performed in duplicate carrying out independent measurements using whole blood obtained from two different donors, collected at different time points. Reproducibility between duplicate independent measurements was assessed, and in a few isolated instances in which the samples exhibited greater than 20% variation (calculated based on fold difference percentage [FD%] rather than absolute CT values), a third independent measurement was acquired. Of the three resulting values, the two closest replicates demonstrating a variation of less than 20% were selected, and their mean fractional difference was used for statistical evaluation.

The assay primarily evaluates early clot initiation and propagation under static conditions. Furthermore, CT reflects clot initiation dynamics and cannot distinguish between intrinsic, extrinsic, or contact pathway activation in the absence of complementary mechanistic assays.

**3. Sample collection and preparation, C3a ELISA, and statistical analysis** are provided in Supplementary Material and Methods

## Results

### 1. Principles and development of thromboelastometry of complement-driven immunothrombosis (TCDI)

The principle of the method is based on the capacity of patient’s plasma to activate C3 in whole blood, leading to coagulation alterations^13^. Since whole blood from healthy subjects constitutes a source of complement, mixing it with patient plasma could activate complement and mimic complement-mediated immunothrombosis.

We hypothesized that pharmacological C3 inhibition in donor blood during exposure to patient plasma, using the Cp40 peptide, would suppress clot formation. The assay measures clotting time (CT) in healthy donor whole blood following incubation with patient plasma, in the presence or absence of the C3 inhibitor Cp40. Complement involvement was functionally defined as CT prolongation following C3 inhibition.

Indeed, incubation of healthy donor citrated whole blood with plasma derived from patients with COVID-19 induced a marked prolongation of CT following Cp40 treatment, indicating that clot initiation was partially dependent on complement activation. Based on these observations, we established thromboelastometry of complement-driven immunothrombosis (TCDI) as a functional assay that quantifies complement-dependent immunothrombotic activity through CT prolongation following complement inhibition.

To minimize variability due to multiple donors, CT prolongation following Cp40 treatment was expressed as fractional difference percentage (FD%), representing the percentage change in CT after complement inhibition (CT__Cp40_) compared to untreated conditions (CT__no Cp40_). FD% was calculated using the formula:

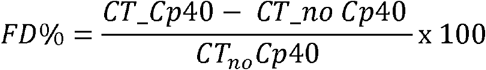

Negative FD% values were assigned a value of 0, indicating the absence of detectable complement-dependent CT prolongation (Figure **1A**).

**Figure 1.**
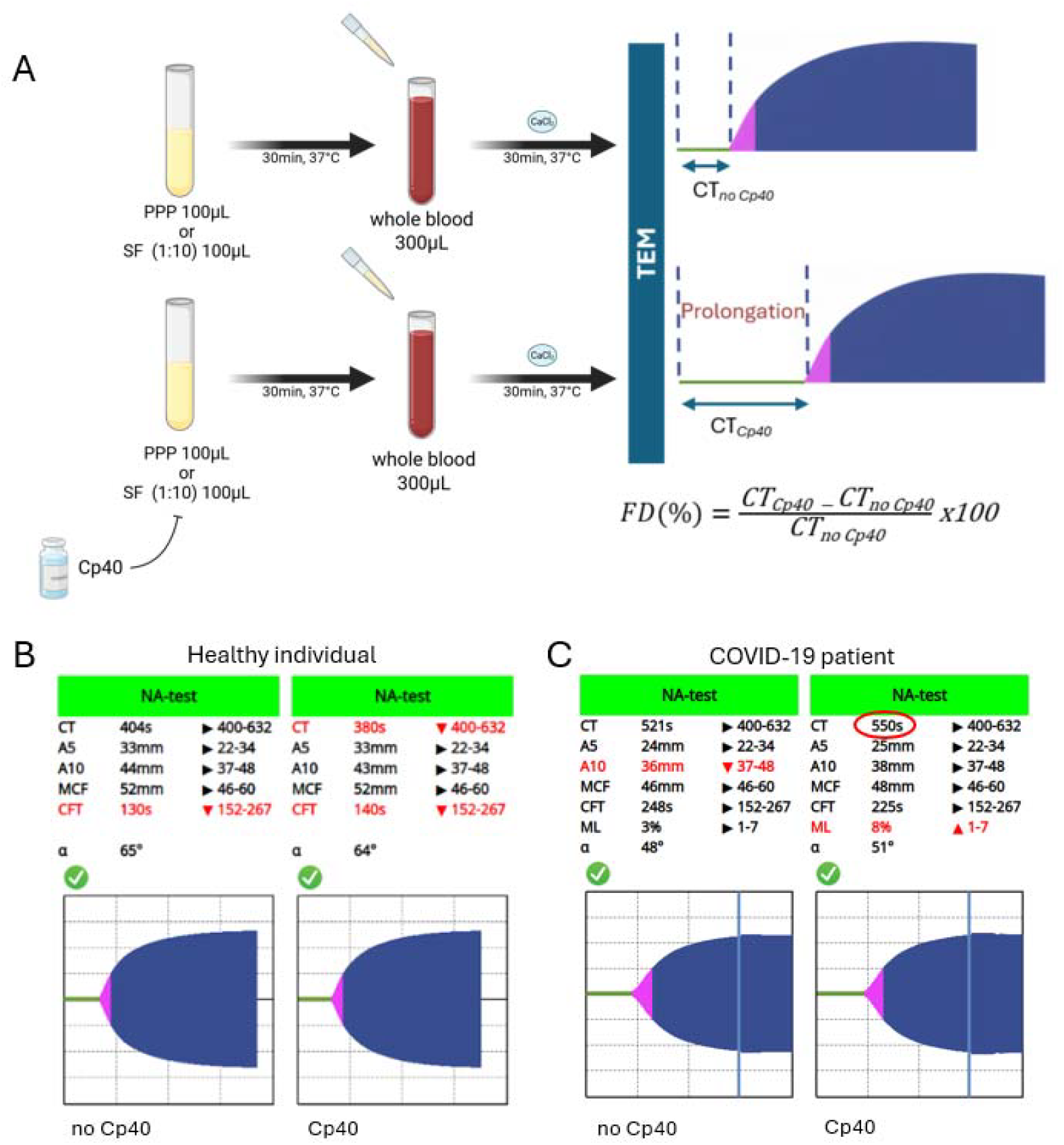
TCDI assay workflow and representative TEM profiles. Healthy donor whole blood was incubated with patient-derived plasma (platelet-poor plasma, PPP) or synovial fluid (SF; diluted 1:10) for 30 min at 37°C, ± Cp40 (pre-incubated with PPP/SF). CaCl_2_ was then added to samples and clot formation was assessed by thromboelastometry (TEM). Clotting time (CT) was the primary readout. FD% was calculated as (CT_Cp40_ – CT_no Cp40_) / CT_no Cp40_ × 100. (**B**) Representative TEM profiles showing the effect of complement inhibition with the C3 inhibitor Cp40 in platelet-poor plasma (PPP) from a healthy individual, compared with the untreated condition. **(C)**Representative TEM profiles demonstrating the effect of Cp40-mediated complement inhibition in PPP from one representative Covid-19 patient PPP sample. PPP: Platelet poor plasma; TEM: Thromboelastometry, TCDI: thromboelastometry of complement-driven immunothrombosis. Created using BioRender.com.

A schematic overview of the TCDI workflow is shown in Figure **1A**, while representative thromboelastometry profiles from healthy donors and patients with COVID-19 are presented in Figures **1B-C**.

### 2. Study of TCDI in clinical diseases driven by immunothrombosis

Compared to healthy donors (Fig. **2A**), APS samples demonstrated significant CT prolongation following Cp40 treatment, indicating active complement-driven immunothrombosis (Fig. **2B**). Consistently, FD% values were significantly increased in APS patients (Fig. **2C**), reflecting enhanced complement-dependent coagulation activity. Notably, two out of six APS patients showed normalization of CT values 3 years later, coinciding with clinical remission and loss of previously detectable antibodies, suggesting a connection with disease serological activity (**Table 2**).

**Figure 2.**
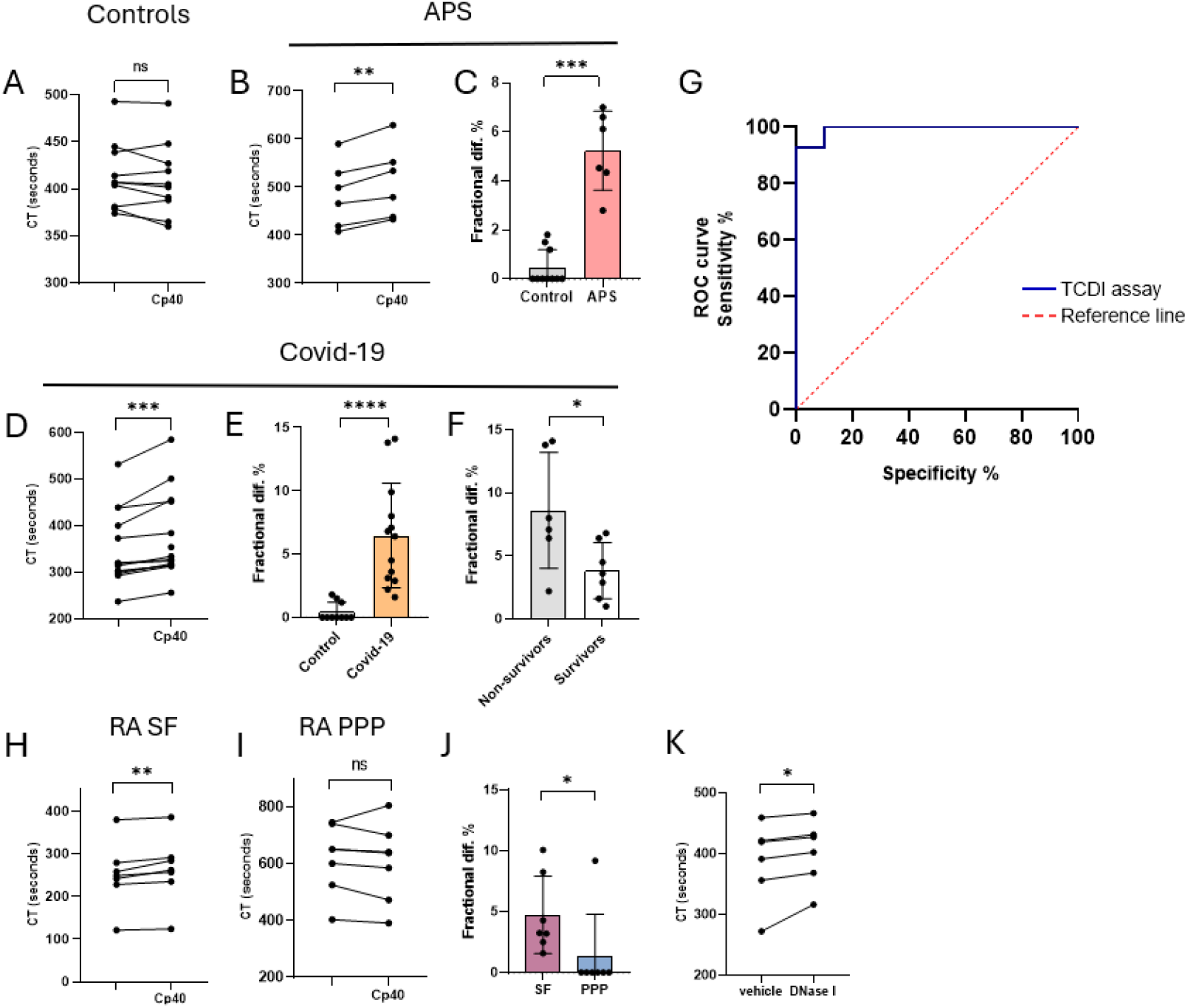
Assessment of complement-driven immunothrombosis in systemic and local inflammatory conditions using the thromboelastometry of complement-driven immunothrombosis (TCDI) assay. (**A–B**) Clotting time (CT; seconds) measured in healthy donor whole blood ± Cp40 following addition of plasma from healthy controls (A; n=10) or APS patients (B; n=6). (**C**) Fractional difference (FD%) comparison between control and APS samples shown in panels A and B. (**D**) CT measured under the same conditions using plasma from severe COVID-19 patients (n=13). (**E**) FD% comparison between control and COVID-19 samples shown in panels A and D. (**F**) TCDI values (FD%) in COVID-19 patients stratified by clinical outcome (Non-survivors vs Survivors). (**G**) Receiver operating characteristic (ROC) curve of the TCDI assay. A cutoff of 2 (values >2 defined as TCDI-positive) provided optimal discrimination between patients and healthy controls, with 92.86% sensitivity and 100% specificity. (**H-I**) CT measured following addition of RA SF (H) or PPP (I). (**J**) Fractional difference (FD%) comparison between RA SF and PPP samples shown in panels H and I (**K**) Effect of NET dismantling (DNase I) on clot formation: CT quantification (n=6). Statistical analysis: Panels (A), (B), (I), and (K) were analyzed using a paired two-tailed Student’s t-test. Panels (C) and (E) were analyzed using the Mann–Whitney test. Panel (D) and (J) were analyzed using the Wilcoxon matched-pairs signed-rank test. Panels (F) and (H) were analyzed using an unpaired two-tailed Student’s t-test. Panel (L) was analyzed using nonparametric Spearman correlation. *p < 0.05, **p < 0.01, ***p < 0.001; ****p < 0.0001; ns, not significant. APS, antiphospholipid syndrome; NET, neutrophil extracellular traps; PPP, platelet-poor plasma; RA, rheumatoid arthritis; SF, synovial fluid; TCDI: thromboelastometry of complement-driven immunothrombosis

Similarly, plasma from severe COVID-19 patients induced significant CT prolongation following Cp40 treatment (Fig. **2D**), with increased FD% compared to controls (Fig. **2E**), supporting C3-driven immunothrombotic activity. Importantly, higher TCDI levels were associated with mortality (Fig. **2F**).

Receiver operating characteristic (ROC) curve analysis of FD% effectively distinguished patient from healthy control PPP samples (Fig. **2G**, **Supplementary Table 2**). An optimal cutoff value of 2 was determined, whereby FD% values greater than 2 were considered TCDI-positive, yielding 92.86% sensitivity and 100% specificity.

Among 7 RA patients in whom PPP and SF were collected simultaneously, significant complement-driven CT prolongation was observed in SF samples (Fig. **2H**), whereas no significant response was detected in peripheral plasma (Fig. **2I**, **Table 3)**. The significant increase of FD% in SF compared with PPP in RA patients (Fig. **2J**), suggests localized complement-driven immunothrombosis within the inflamed joint compartment. Notably, one patient exhibited a positive TCDI signal in both compartments, indicating a more pronounced systemic and local immunothrombotic phenotype within the RA group.

Given that neutrophil extracellular traps (NETs) are involved in the immunothrombotic pathway downstream of activated C3, in COVID-19 patients^5^ we assessed their contribution to plasma-induced clot formation. Patient PPP samples were pre-treated with DNase I alone to dismantle NET structures prior to incubation with donor whole blood. DNase I treatment significantly prolonged CT compared with untreated conditions **(Fig. 2K**). Notably, the pattern of CT prolongation observed following DNase I treatment was similar to that induced by Cp40, in accordance with findings suggesting a functional interplay between complement activation and NET-mediated coagulation.

### 3. The TCDI positivity is correlated with higher levels of C3a generated upon stimulation

Plasma isolated in parallel from the same PPP/whole blood mixtures used for TEM analysis was also analyzed for C3a generation by ELISA in both conditions, i.e. the absence and presence of Cp40 treatment. C3a levels were significantly higher in TCDI-positive compared with TCDI-negative samples (Fig. **3A**). In TCDI-positive samples, treatment with Cp40 markedly reduced C3a levels (Fig. **3B**), whereas a relatively modest reduction was observed in TCDI-negative samples (Fig. **3C**). Accordingly, ΔC3a, defined as the Cp40-mediated reduction in C3a levels (C3a _noCp40_-C3a _Cp40_), was significantly greater in TCDI-positive than in TCDI-negative samples (Fig. **3D**).

**Figure 3.**
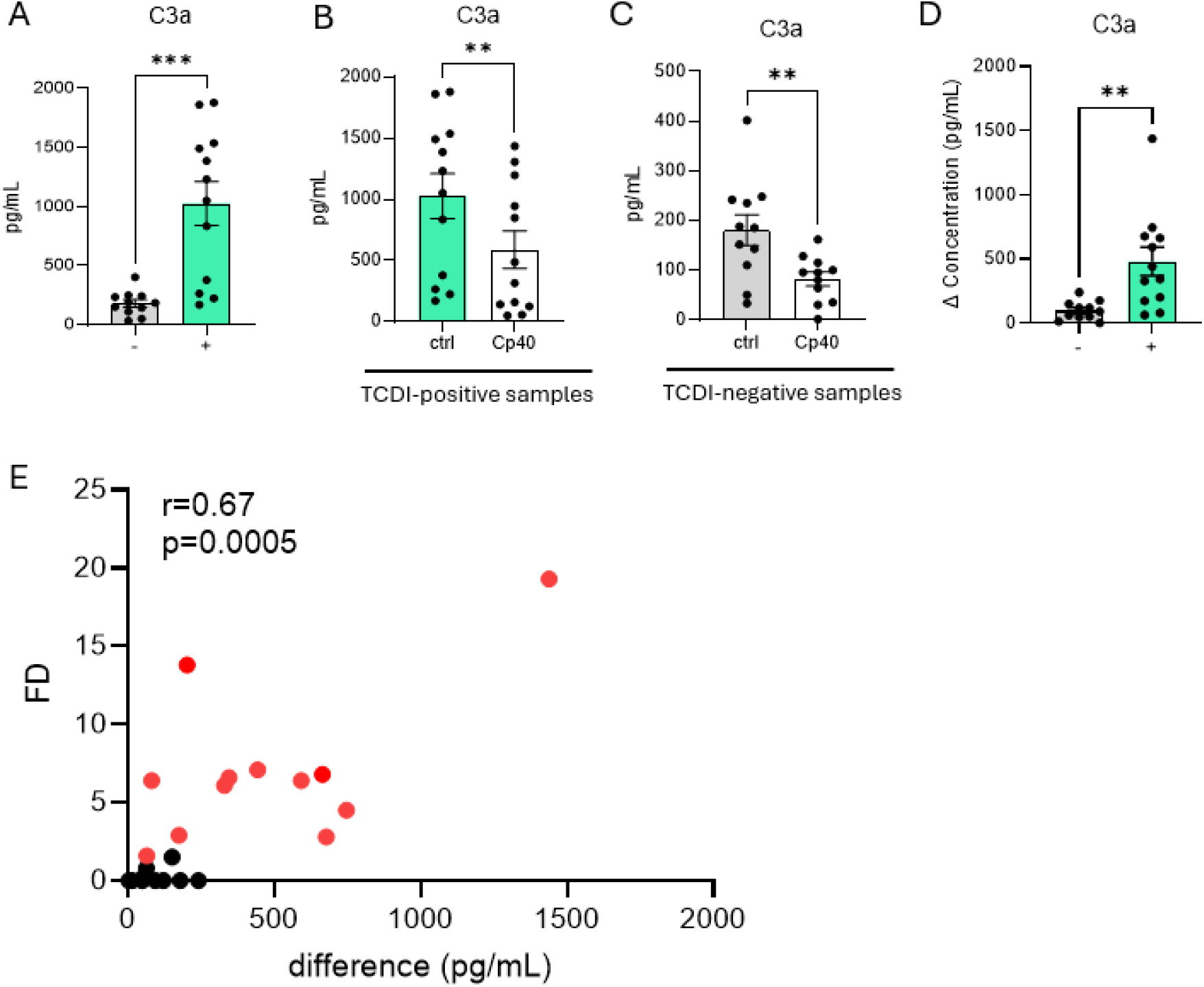
(**A**) Plasma C3a levels (ELISA) following incubation of whole blood with TCDI-negative or TCDI-positive samples. (**B)** Plasma C3a levels (ELISA) following incubation of whole blood with TCDI-positive samples in both conditions, absence and presence of Cp40 (**C**) TCDI-negative samples in experimental conditions as in B. (**D)** Differences in C3a levels (ΔC3a) after Cp40 treatment in TCDI-negative and TCDI-positive samples. (**E**) Correlation between TCDI (FD%) and Cp40-mediated differences in C3a ELISA levels (ΔC3a; pg/mL). Panels (A) and (D) were analyzed using an unpaired parametric t-test. Panels (B) and (C) were analyzed using the Wilcoxon matched-pairs signed-rank test. Panel (D) was analyzed using an unpaired parametric t-test. Panel (E) was analyzed using nonparametric Spearman correlation. *p < 0.05, **p < 0.01, ***p < 0.001; ****p < 0.0001; ns, not significant. TCDI: thromboelastometry of complement-driven immunothrombosis; FD, Fractional difference

Since Cp40 inhibits C3 activation by acting on nascent C3 convertases and on native C3 made available as a substrate from the freshly added whole blood, it does not affect pre-existing C3a levels present in the samples but only prevents the generation of newly formed C3a during the assay. Therefore, to account for baseline C3a levels and specifically assess newly generated C3a, we correlated the Cp40-mediated difference in C3a levels (ΔC3a) with the corresponding TCDI values (FD%) of each sample, (Fig. **3E**). A significant positive correlation was observed between ΔC3a and FD% (r = 0.67, p = 0.0005; Fig. **3E**), indicating that the extent of C3 activation that can be prevented by Cp40 treatment is closely associated with complement-driven immunothrombosis. These findings further support the biological relevance of TCDI as a functional measure of complement-mediated immunothrombotic activity.

## Discussion

The TCDI assay presented here is based on the capacity of a patient’s plasma to activate the complement protein C3 when mixed with healthy donor whole blood, thereby inducing measurable alterations in coagulation dynamics. By selectively inhibiting C3 activation with Cp40, the assay determines the relative contribution of complement activation to the thrombotic response. This contribution is quantified by the fractional difference percentage (FD%) in clotting time, providing a standardized and functional measure of complement-dependent immunothrombosis.

This methodology addresses a major unmet need in current diagnostics. While the mechanistic relationship between complement activation and coagulation has been extensively studied, functional tools capable of dynamically measuring this interaction and capturing the contribution of its separate components in clinical samples remain limited. To our knowledge, this represents the first functional assay capable of visualizing and quantifying complement-driven immunothrombosis in real time.

By directly measuring the contribution of complement activation to clot formation, TCDI could help identify patients in whom complement is an active driver of immunothrombotic disease rather than a bystander mechanism or secondary biomarker. This could support patient stratification, comprehensive disease monitoring, and the rational selection of complement- or coagulation-targeted therapies.

In the context of severe COVID-19, the TCDI assay served as a functional indicator of immunothrombotic activity that correlated directly with clinical outcome. Our data reveal a significant association between higher TCDI levels and mortality. These findings strongly imply that the magnitude of complement-mediated immunothrombosis acts as a critical determinant of both disease severity and overall patient survival ^14^. They also complement the clinical and biological insights gained from the Phase II evaluation of the C3 inhibitor AMY-101 in severe COVID-19, further supporting the therapeutic relevance of C3-targeted intervention in severe immunothrombotic diseases.

Our findings in APS further highlight the potential sensitivity of the assay to disease activity. Samples obtained from patients with active APS exhibited a pronounced complement-dependent signal as previously described^15–17^. Notably, in two patients reassessed three years later, TCDI values were normalized in parallel with clinical remission and the loss of previously detectable antiphospholipid antibodies (**Table 2**). Although limited by the small sample size, this observation suggests a potential association between TCDI levels and serological disease activity.

In RA, TCDI positivity was predominantly detected in SF rather than peripheral plasma, demonstrating that complement-driven immunothrombosis is largely confined to the local joint microenvironment, consistent with recent observations in RA^18^. These findings highlight the ability of the assay to distinguish localized from systemic immunothrombotic activation and may have important implications for selecting the most appropriate route of administration for complement or coagulation inhibitors, thereby maximizing therapeutic benefit in autoimmune and inflammatory diseases such as RA.

Several technical optimizations were essential to simplify the application of the assay. According to the principle and the design of TCDI, derivatives of dicoumarol or low molecular weight heparin (LMWH), do not interfere with TCDI measurements. This concept aligns with observations made during optimization. No changes in the pattern of TCDI values were observed in samples from the same patient drawn before and after initiation of anticoagulation therapy with dicoumarol or heparin.

A critical finding of our study was the strong positive correlation between TCDI values (FD%) and the amount of newly generated C3a during the assay. In TCDI-positive samples, C3a levels were significantly higher compared to negative samples, and this elevation was markedly reduced by Cp40 treatment. Since Cp40 inhibits C3 activation but does not affect pre-existing C3a, this correlation supports that the CT prolongation is a direct functional reflection of complement activation.

Beyond its diagnostic potential, TCDI may also serve as a functional assay for future mechanistic and therapeutic studies. The assay could be used to evaluate complement-targeted therapies, investigate disease-specific immunothrombotic signatures, monitor treatment responses, and identify patient subsets with heightened complement-driven immunothrombotic risk. Further studies involving larger patient cohorts are required to validate the clinical applicability of this assay across diverse disease phenotypes associated with complement-mediated immunothrombosis. Additional technical improvements may further enhance its translational utility.

## Data availability statement

The data that support the findings of this study are available from the corresponding author upon reasonable request.

## Ethics approval and patient consent statement

The study was approved by the Ethics Committee of University Hospital of Alexandroupolis (approval number: 10769) and conducted in accordance with the Declaration of Helsinki. All participants had previously provided written informed consent for the use of their biological samples for research purposes.

