## Supplementary Material for "A Modified Thromboelastometry Assay Enables Rapid, Real-Time Evaluation of Complement-Driven Immunothrombosis"

**Supplementary materials and methods**

**1. Collection of platelet-poor plasma (PPP) and synovial fluid (SF)**

Venous blood from healthy individuals (HI) was collected into sodium citrate anticoagulant tubes (BD Vacutainer®, Becton, Dickinson and Company). Samples were centrifuged at 500 × g for 15 minutes at room temperature to obtain PPP. The supernatant plasma was carefully collected to avoid platelet contamination, aliquoted, and stored at −80°C until further analysis.

SF samples were obtained from patients undergoing arthrocentesis. Immediately after collection, samples were transferred into sodium citrate vacutainer tubes. SF was centrifuged at 800 × g for 10 min at 4 °C to remove cells and debris. The resulting supernatant was either used immediately for experiments or aliquoted and stored at –80 °C until further analysis.

**2. C3a ELISA**

Following incubation under identical experimental conditions, PPP/whole blood mixtures were either subjected to TEM analysis or centrifuged at 500 × g for 15 min at room temperature to isolate plasma for subsequent measurement of C3a levels by ELISA. Due to limited availability of biological material, C3a measurements were performed in a subset of samples. C3a levels were quantified using a commercially available human C3a ELISA kit (HK354; Hycult Biotech, Uden, the Netherlands), according to the manufacturer’s instructions. Samples were analyzed in duplicate and, where applicable, measured in parallel with or without Cp40 treatment.

**3. Statistical analysis**

Statistical analysis was performed using GraphPad Prism software (version 9.0; San Diego, CA, USA). Data are expressed as mean ± standard error of the mean (SEM). Two-group comparisons were analyzed using a paired, two-tailed t-test when the data were normally distributed. For paired data that did not follow a normal distribution, the Wilcoxon signed-rank test was employed. The receiver operating characteristic (ROC) curve was performed to illustrate the performance of varying threshold settings; the selected cut-off corresponded to the maximum Youden's J value. Statistical significance was defined as *p<0.05, **p<0.01, ***p<0.001, ****p<0.0001.

**Supplementary Tables**

| **Patient ID** | **Age** | **Gender** | **FD** |
| --- | --- | --- | --- |
| **HI 1** | 20-29 | F | 0 |
| **†HI 2** | 30-39 | F | 0 |
| **†HI 3** | 20-29 | M | 1.2 |
| **HI 4** | 20-29 | M | 0 |
| **†HI 5** | 20-29 | F | 1.8 |
| **†HI 6** | 30-31 | M | 0 |
| **HI 7** | 20-29 | M | 0 |
| **†HI 8** | 40-49 | M | 0 |
| **†HI 9** | 20-29 | F | 0 |
| **†HI 10** | 20-29 | F | 1.5 |

***Supplementary Table 1****. Healthy blood donor characteristics. HI, healthy individual; FD, Fractional Difference; M, Male; F, Female*

***†*** *These samples were available for C3a ELISA.*

| **TCDI cutoff** | **Sensitivity (%)** | **Specificity (%)** | **Youden's J** |
| --- | --- | --- | --- |
| > 0.60 | 100 | 70 | 0.7 |
| > 1.35 | 100 | 80 | 0.8 |
| > 1.55 | 100 | 90 | 0.9 |
| > 1.70 | 92.86 | 90 | 0.8286 |
| **> 2.00** | **92.86** | **100** | **0.9286** |
| > 2.55 | 85.71 | 100 | 0.8571 |
| > 3.00 | 78.57 | 100 | 0.7857 |

***Supplementary Table 2:*** *TCDI-negative versus TCDI-positive (COVID-19) discrimination using ROC analysis of FD%: Sensitivity and specificity of proposed cutoff values; Youden’s J is maximized at >2.000, yielding 92.86% sensitivity and 100% specificity. TCDI: thromboelastometry of complement-driven immunothrombosis.*
